# Uveal and cutaneous melanoma share a common mutation with distinct prognostic implications: A bioinformatic study

**DOI:** 10.64898/2026.08.07.26359988

**Authors:** Fatemeh Razmjooei, Hamidreza Ashayeri, Zahra Jafarzadeh, Porya Dabbaghabdollahi, Ali Jafarizadeh

## Abstract

**What was known before:**

1. Uveal melanoma (UM) and cutaneous melanoma (CM) arise from a common melanocytic lineage but exhibit distinct genetic landscapes, biological behavior, and clinical outcomes.
2. Pigmentation-related genes, including IRF4 and HERC2, have been implicated in melanoma susceptibility and prognosis; however, the extent of shared genetic variants between UM and CM has remained unclear.
3. Previous studies have primarily investigated UM and CM separately, leaving limited understanding of their common genetic architecture and potential shared prognostic biomarkers.

**What this study adds:**

1. This bioinformatic analysis identified only two shared validated genetic variants between UM and CM across publicly available databases: rs12203592 (IRF4) and rs12913832 (HERC2).
2. Protein–protein interaction analysis demonstrated that IRF4 and OCA2 directly interact with HERC2, supporting a biologically plausible pigmentation-related pathway linking the two melanoma subtypes.
3. The findings suggest that IRF4 may represent a common prognostic biomarker across both UM and CM, while combined assessment of IRF4 and HERC2 could improve future risk stratification and facilitate more personalized therapeutic approaches.

**Background:** Uveal melanoma (UM) and cutaneous melanoma (CM) both originate from the same cell line. This proposes the possibility of a shared mechanism between entities, requiring explicit investigation.

**Methods:** Data from GWAS Catalog and DisGeNET were used to identify shared variation-disease associations (VDAs) between UM and CM. The results were validated using the Ensembl database. In the next step, the STRING database was used to identify the protein– protein interaction.

**Results:** Subsequently, 109 unique VDAs were identified for UM and 880 for CM. However, only 2 VDAs were found to be shared among UM and CM in different ethnic groups. These shared VDAs were rs12203592 of the IRF4 gene, rs12913832 of the HECT and RLD domain-containing E3 ubiquitin protein ligase 2 (HERC2) gene. Notably, PPI network assessment through STRING showcased that OCA2 and IRF4 directly interacted with HERC2.

**Conclusion:** While HERC2 acts as a poor prognostic factor in uveal melanoma, IRF4 status is a key prognostic indicator in both UM and CM. Identifying IRF4 allele contributions enables a better understanding of melanoma pathogenesis and fosters the development of disease-specific approaches.

## 1. Introduction

Melanocytes are pigment-producing cells derived from the neural crest and can be found in both uveal and skin tissue [1]. Melanoma results from the malignant transformation of melanocytes[2]. Cutaneous melanoma (CM) is the most common type of melanoma; however, melanoma can occur in other tissues, such as uveal melanoma (UM) [3]. Although the CM and UM may seem to originate from the same cell line, the pathophysiology and clinical outcomes differ significantly [1]. For instance, ultraviolet light (UV) and related genetic mutations are a main risk factor for CM development, while probably not affecting UM development [4].

It is considered that the main driver mutations between UM and CM differ, despite having the same origin [5, 6]; however, they both share a common risk factor. Fair skin is reported to increase the risk of both UM and CM [3]. This suggests that genes involved in pigment production may play a part in both conditions, either directly or indirectly, and independent of the UV light [4]. Melanin acts as an antioxidant in melanocytes, and mutations in this pathway may decrease its protective effects [7]. Mutations can also act by reducing transcription factors (TF), leading to a simultaneous change in the expression of multiple genes [8]. For example, interferon regulatory factor-4 (IRF4) acts as a TF, involved in pigmentation and anti-tumor activity [9].

On the other hand, the HECT and RLD domain-containing E3 ubiquitin protein ligase 2 (HERC2) gene targets key DNA damage response proteins via the ubiquitin–proteasome system [10]. The HERC2-associated variant was shown to be the main determinant of blue eye color in European ethnicity, and also contributes as a risk factor for cutaneous melanoma [11], specifically in individuals with a low-risk phenotype such as pigmented skin [12]. This implies a possibility of a shared mainstream pathway between UM and CM, which may play a role alongside the main mutation in each entity.

Bioinformatics methods are useful tools for leveraging biomedical data and understanding gene-disease associations [13]. Identifying the genetic profile of malignancies is associated with improved prognostic prediction and therapeutic targets. Since CM and UM share a common cell lineage, the same genetic mutations may be present. In this study, we used an integrative bioinformatics pipeline to systematically compare the genetic landscapes of CM and UM, uncovering genetic similarities. These similarities can provide insights into the pathophysiology of both conditions, leading to enhanced diagnosis and treatment strategies.

## 2. Materials and Methods

Human-based genes and variation-disease associations (VDAs) were found using the GWAS Catalog [14] and DisGeNET (https://www.disgenet.com) [15]. The search in all databases was updated on June 06, 2026. The GWAS catalogue (v1.0.3-genome-wide significant threshold of 10⁻^6^), and DisGeNET (v7.0) were searched using disease terms such as: “melanoma”, “melanocytoma”, “cutaneous melanoma”, “cutaneous melanocytoma”, “uveal melanoma”, “uveal melanocytoma”, “malignant melanoma”, “advanced cutaneous melanoma”, “uveal epithelioid cell melanoma”, “advanced uveal melanoma “. To avoid misleading differences across variant categories in databases, we harmonized the GWAS catalogue (common variants) and DisGeNET (text-mined variants) using gene symbols (HGNC) and rsIDs. Human studies and variants with available p-values were included. Animal studies and phenotypes unrelated to cutaneous and uveal melanoma were excluded.

Afterward, the genes and VDAs associated with CM and UM were downloaded and compared using the Molbiotools comparator (https://www.molbiotools.com). Then, the shared genes and VDAs were checked in the Ensembl database [16]. Ensembl assessed the presence of the reported VDAs in the original research, the statistical significance of the reported association, and the consistency of the findings across different sources and studies. Variants meeting the following criteria were ‘Validated’ variants: the association was reported in a GWAS publication indexed in Ensembl, p-values were reliable, and findings were consistent across two or more sources. Conflicting variants were omitted from the analysis. The presence, significance, and consistency of reported VDAs were evaluated independently by two authors. Any disagreements and discrepancies were resolved through discussion and consultation with the third author.

The Human Protein Atlas (HPA) database (v21.5) was used to assess protein and mRNA expression of shared VDA-related genes and to identify their potential functions in the eye and skin [17]. Protein–protein interaction (PPI) networks were constructed using the STRING v12. Database using a confidence score threshold of 0.7 [18]. Gene lists of interest were submitted to STRING, and only “protein–protein interactions” for human proteins were considered. We used the “multiple sources” option, integrating evidence from experimental data, curated databases, co-expression, and text mining, as provided by STRING. To reduce noise and focus on biologically relevant interactions, we applied a minimum required interaction score of 0.7, corresponding to high-confidence edges.

This study did not involve any cell lines or in vitro experimentation. All analyses were performed using publicly available databases and bioinformatics tools; therefore, cell line authentication by an expert and mycoplasma contamination testing were not applicable.

## 3. Results

In a search across two databases, the GWAS catalogue and DisGeNET, 109 unique VDAs were identified for UM and 880 for CM. However, only 2 VDAs were found to be shared among UM and CM in different ethnic groups. These shared VDAs were rs12203592 of the IRF4 gene, rs12913832 of the HECT and RLD domain-containing E3 ubiquitin protein ligase 2 (HERC2) gene, as depicted in **Fig 1**. A complete list of VDAs for each disease is available in S1 Table.

**Fig 1.**
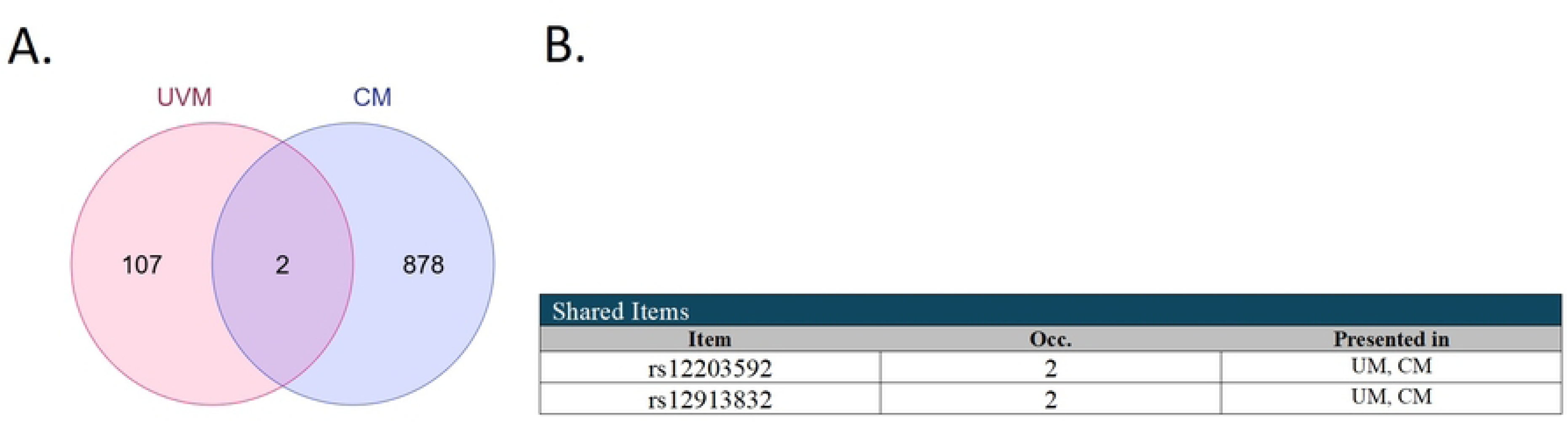
Venn diagram (A.) and intersection table (B.) of variation disease associations (VDAs) showing the shared number of VDAs between uveal and cutaneous melanoma. CM: Cutaneous melanoma, UM: Uveal melanoma.

To evaluate mRNA and protein expression among various tissues, IRF4 and HERC2 were investigated through the HPA. In the skin tissue, only IRF4 mRNA expression was evident, while data on IRF4 mRNA and protein expression in the eye were sparse. In addition, although both mRNA and protein expression of HERC2 were detected in the skin, only mRNA expression was reported in the eye. Their distribution in these organs highlights their potential functional coalescence with rising debates about differences in expression. **Fig 2** represents the IRF4 and HERC2 mRNA and protein expression in various tissues.

**Fig 2.**
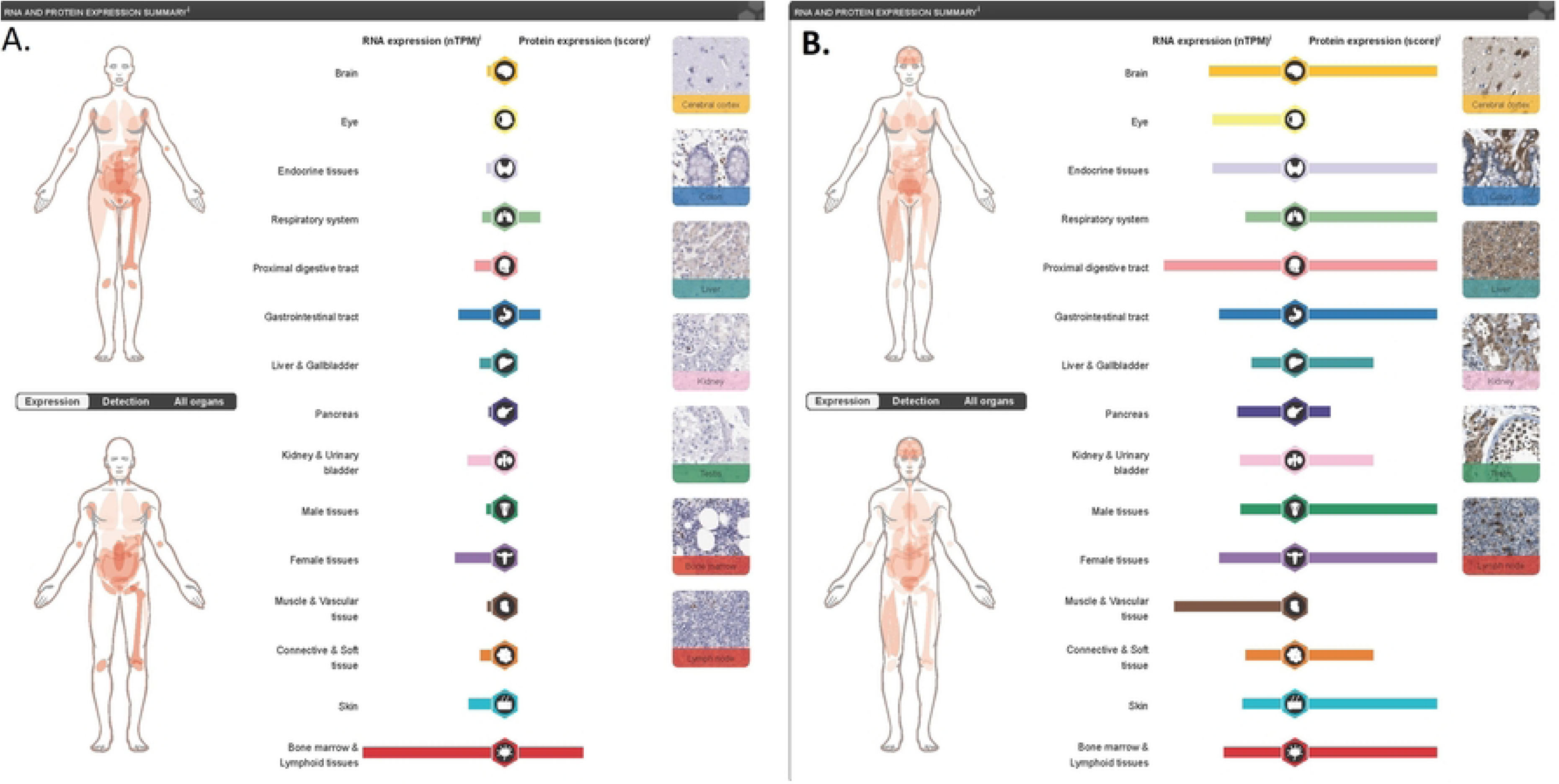
Graphical data from the Human Protein Atlas (HPA) showcases the IRF4 (A.) and HERC2 (B.) mRNA and protein expression.

Subsequently, the interaction PPI network was constructed. The resulting network included 12 interconnected nodes: Ubiquitin-specific Protease-35 (USP35), USP16, myelin transcription factor-1 (MYT1), HERC2, F-Box and Leucine Rich Repeat Protein −5 (FBXL5), Dedicator Of Cytokinesis-10 (DOCK10), Neuralized-like protein-4 (NEURL4), Rab geranylgeranyltransferase (RABGGTA), Oculocutaneous albinism type II (OCA2), IRF4, Spi-B Transcription Factor (SPIB), and Guanine Nucleotide Exchange Factor (DEF6). Notably, OCA2 and IRF4, as established regulators of pigmentation, directly interacted with HERC2, suggesting a possible physical link between melanosome function and ubiquitin pathways. **Fig 3** represents the PPI network.

**Fig 3.**
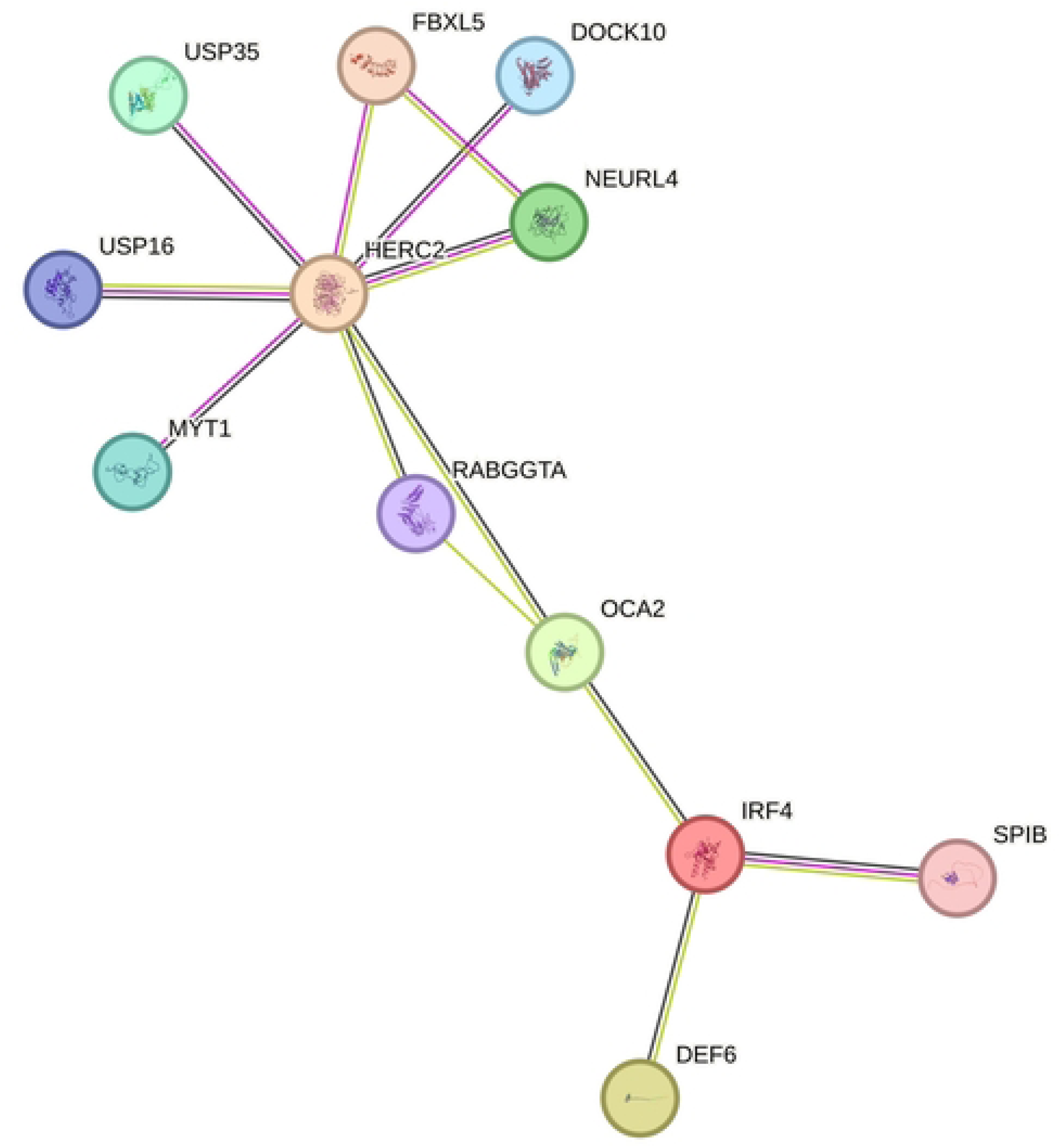
STRING illustrated the protein-protein interaction network for IRF4 and HERC2.

## 4. Discussion

In this study, we identified two common single-nucleotide polymorphisms (SNPs), namely, rs12203592 and rs12913832 in the IRF4 and HERC2 genes, respectively, that are shared between CM and UM. The IRF4 gene is located on chromosome 6, and the reported SNP is in position 6:396320 [19]. The rs12913832 SNP is in intron 86 of the HERC2 gene and is believed to have an impact on OCA2 expression [20].

The rs12913832 can influence the OCA2 expression and the pigment production in melanocytes, which affects eye and skin color [21]. Ferguson et al. report that alleles associated with higher OCA2 expression (darker eyes) decrease the odds of UM [19]. The G/G homozygote allele is associated with poor UM outcome due to the high presence of monosomy 3 [22]. These results may be due to the melanin content of cells. The darker colors contain more eumelanin than lighter colors; in return, light skin has higher pheomelanin content [23]. Eumelanin has anti-oxidative properties, while pheomelanin has more potential to create a pro-oxidant environment [22, 24, 25].

Studies show contradictory results regarding the association between rs12913832 and CM [26, 27]. In another study, the rs12913832 shows association with CM development with a p-value of 0.042; however, it failed to reproduce the results in patients with 100% Northern European ancestry (p-value: 0.51) [28]. One study reported the pooled odds ratio of CM development for the G-allele based on data from the MD Anderson Study, Harvard Nurse Health Study, and the Harvard Health Professionals Follow-Up Study to be 1.29 (95% CI: 1.12-1.48) [29].

Chromosomal abnormalities are more frequently seen in UM than in CM. The HERC2 SNPs are often seen in UM with chromosomal abnormalities. However, it is important to note that they do not add prognostic value when the chromosome 3 monosomy is present [30]. While in CM, chromosomal abnormalities are rarely seen [31], which may reduce the importance of rs12913832 in their development and prognosis.

The rs12203592 SNP in IRF4 has two main reported alleles. The rs12203592*C allele is the major allele, while the rs12203592*T allele is the minor allele located in intron 4 of the IRF4 gene [32]. In both uveal and skin tissues, the T allele was associated with reduced IRF4 expression by preventing the TFAP2α binding [33]. According to the current literature, IRF SNPs manifest different behaviors in melanoma development based on their location.

The T allele of IRF4 was reported to increase the odds of UM [34, 35]. However, the data about the association of these alleles with CM show conflicting results. One study also found that the major allele is related to the truncal CM in Australia, the UK, and Sweden [36]. Another study reported protective effects of the T allele in truncal CM, while showing increased mortality in head and neck CM patients with the same SNP [37]. Development of truncal CM has a stronger association with a higher nevus count [38], which in adults is more commonly seen with the C allele [28], while the development of head and neck CM has a stronger association with actinic keratosis [38], which is more commonly seen in the T allele [39, 40].

Also, IRF’s status has complex effects on the patient’s prognosis. In UM, the presence of the minor IRF4 allele was associated with better survival, due to its association with disomy of chromosome 3 [41]. Studies performed on US residents, European Americans, and Caucasians residing in the US report the association of the T allele with increased CM development and mortality [42, 43]. One study of 3285 CM patients reports that this allele is associated with higher Breslow thickness and the presence of mitoses, leading to a poor prognosis [44]. However, studies conducted in the Spanish population failed to reproduce these results.

It is also important to consider the effect of such mutations on tumor-immune system interactions. IRF4 is also strongly expressed in lymphoid tissue and plays important roles in immune function. Interestingly, the T allele increases IRF4 expression, compared to the C allele [45], leading to an increase in telomerase activity [45, 46]. Therefore, these SNPs can influence tumor-immune system interactions and affect patients’ prognosis (**Fig 4**).

**Fig 4.**
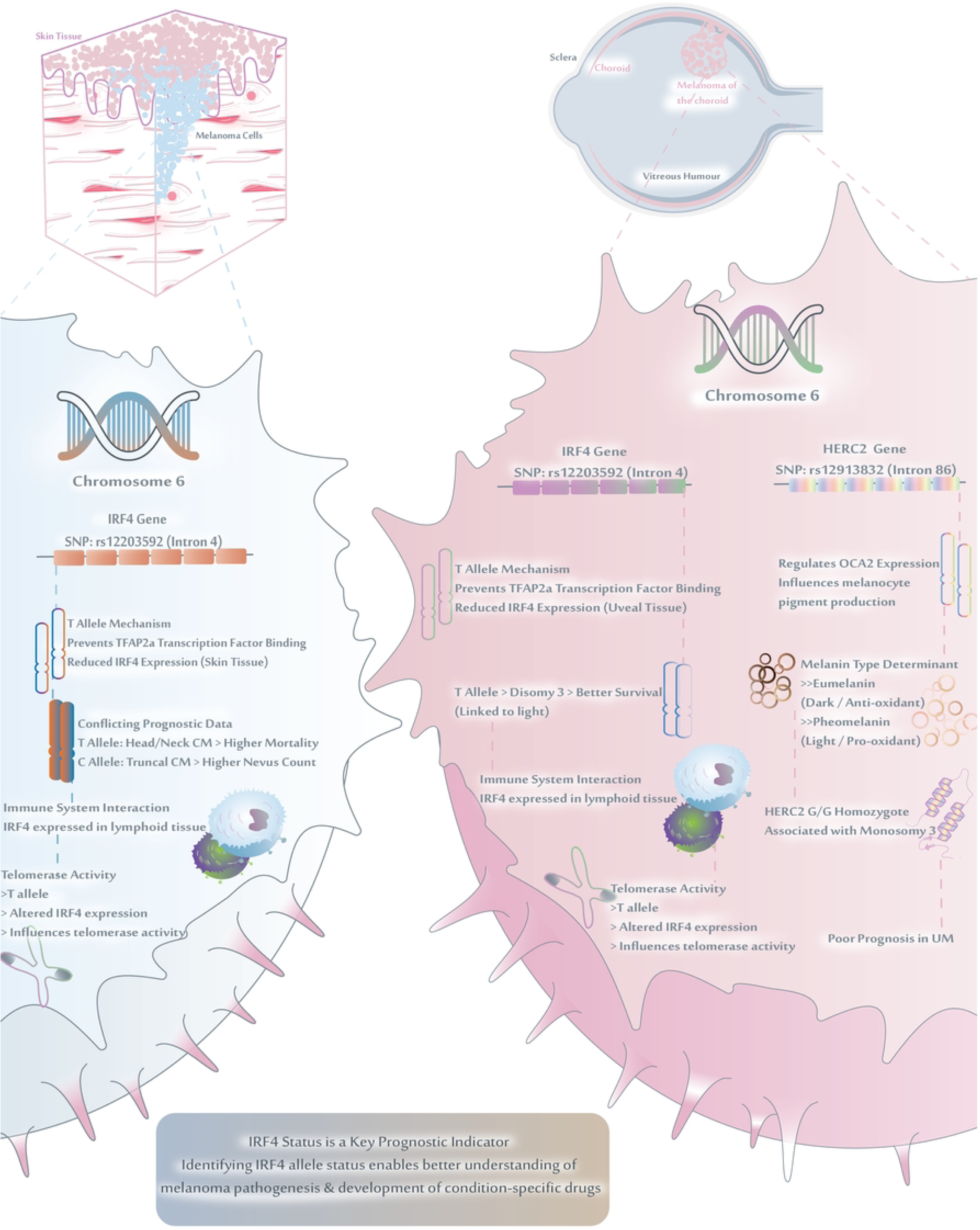
The potential pathways contributing to IRF4 and HERC2 roles in uveal and cutaneous melanoma (Designed by Adobe Illustrator 2026).

Identification of IRF4 VDA as a common factor in both UM and CM provides new insights about the role of these SNPs in disease pathogenesis and prognosis. Identifying IRF4 status can help better understand the risk of melanoma. Targeting the IRF4 gene, considering the patient’s IRF4 allele status, can help develop condition-specific drugs. One important limitation of our study is the reliance on information provided by the databases. These databases are prone to bias, which may affect our results. Other methods, such as in-depth analysis, can identify newer mutations and provide more comprehensive results. We also didn’t evaluate the epigenetic similarities and differences between UM and CM. Identifying these epigenetic factors, such as miRNAs, may provide newer diagnostic and prognostic tools for better diagnosis and management. Further longitudinal studies with larger scales are required to determine the exact role of rs12203592 in each type of melanoma.

## 5. Conclusion

Our study highlights two common variants, rs12203592 (IRF4) and rs12913832 (HERC2), based on online databases. HERC2 affects the prognosis of uveal melanoma in association with monosomy of chromosome 3. IRF4 plays a role in both tumors as a common prognostic determinant and can control pigmentation, immune system changes, and malignant behavior. The relationship between OCA2, IRF4, and HERC2 has not been studied in detail; however, the role of a pigmentation-related pathway is noted. Given these results, IRF4 allele status may be a promising biomarker for risk stratification and prognosis. Further studies to validate these associations are needed. Eventually, combining IRF4 and HERC2 genetic profiles with clinical data may help with more personalized diagnostic and therapeutic strategies for melanoma patients.

## Acknowledgments

The authors used Grammarly to check grammar and improve sentence clarity. All final text was reviewed and edited by human authors, who take full responsibility for the content.

## Ethics statement

This study was conducted using publicly available, de-identified data from established databases (GWAS Catalog, DisGeNET, and Ensembl) and did not involve human participants or identifiable personal data. Therefore, ethics approval and informed consent were not required. The Declaration of Helsinki was strictly followed during the course of this study.

## Data availability statement

The data that support the findings of this study are openly available in GWAS Catalog [at https://www.ebi.ac.uk/gwas/], DisGeNET [at https://disgenet.com/], and Ensembl [at https://www.ensembl.org/index.html].

## Supporting information

**S1 Table. Complete list of variant disease associations (VDA) from DisGeNET and GWAS databases for UM and CM.**

